# Ethnicity, migration, and weight misperception: a Born in Bradford study

**DOI:** 10.1101/2021.07.07.21259906

**Authors:** Tiffany C Yang, Kimberly P Truesdale, Brian Kelly, Marena Ceballos Rasgado, Maria J Bryant

**Affiliations:** Bradford Institute for Health Research, Bradford Teaching Hospitals NHS Foundation Trust, Bradford, BD9 6RJ, United Kingdom; University of North Carolina Gillings School of Global Public Health, Chapel Hill, North Carolina, 27599, United States; Department of Health Sciences, University of York, York, YO10 5DD, United Kingdom; Hull York Medical School, University of York, York, YO10 5DD, United Kingdom

## Abstract

**Background:** Weight perception may differ by ethnicity but is not well-understood, particularly among migrants to the United Kingdom (UK). It is also unknown whether a figure rating scale (FRS) or perceived weight question (PWQ) is more accurate for assessing body weight perception.

**Methods:** At 24-months postpartum, women in the Born in Bradford cohort (Bradford, UK) completed the 9-item Stunkard FRS and a 7-category PWQ. Both scales were condensed to weight categories representing the World Health Organisation cut-offs. Weighted kappa statistics assessed agreement between measured and perceived weight categories. χ^2^ and Fisher’s exact tests assessed categories of misperception by ethnicity and migration and logistic regression was used to examine odds of underestimation.

**Results:** Thirty percent of white British and 23% of Pakistani-origin women had obesity. Agreement between measured and perceived weight categories were similar for white British women using either a visual scale or weight question (FRS: κ [95%CI]: 0.61 [0.56, 0.65]; PWQ: 0.61 [0.55, 0.68)]. Overall agreement was lower for Pakistani-origin women with the visual scale performing better (FRS (0.58 [0.52, 0.63]) vs PWQ (0.47 [0.40, 0.54]). Pakistani-origin women, particularly those born outside the UK, were more likely to underestimate their body weight compared to white British women; this was greater with the PWQ (18% vs 10%; p<0.001) than FRS (14% vs 6%; p<0.001). Pakistani-origin women were 154% more likely to underestimate their body weight compared to white British women with the FRS and 111% more likely to underestimate when using the PWQ.

**Conclusion:** We observed ethnic differences in weight misperception with Pakistani-origin women more likely to underestimate their weight compared to white British women. Our findings suggest visual scales, rather than perceived weight questions, are more appropriate for the self-assessment of body weight.

## Introduction

The epidemic and health consequences of overweight and obesity are well-documented ^(1)^. In the United Kingdom (UK), 58% of women are categorised as having overweight or obesity with higher prevalence among racial and ethnic minority groups ^(2,3)^. Numerous public health campaigns and interventions exist to address excess weight, with many focusing on lifestyle and individual behaviour change ^(4,5)^.

Health promotion messages targeting weight control strategies rest on the assumption that individuals are able to accurately identify their body weight as how individuals perceive their body weight is thought to influence their intentions towards weight change and control ^(6)^. A large proportion of adults fail to correctly identify that they have overweight ^(7,8)^. Research suggests that 31% of women and 55% of men with overweight were unable to accurately perceive their overweight and fewer than 10% of UK obese adults were able to correctly identify themselves as “obese” ^(9,10)^.

The degree of body weight misperception – disagreement between measured and perceived body weight – has been found to differ by ethnicity, and may be due to differences in social norms and cultural beliefs about body size ^(11–15)^. However, the relationship between migration and misperception is not clear for South Asian migrants, one of the largest migrant groups in the UK ^(16–19)^. Additionally, studies of body weight misperception have used either a visual scale or self-perceived weight status question but not both to determine whether one measure is more accurate. The aim of this study was therefore to assess the prevalence of body weight misperception by ethnicity and country of birth using both a visual figure rating scale and a perceived weight status question.

## Materials and methods

### Study population

This study utilised data from the Born in Bradford (BiB) study, a longitudinal birth cohort which aims to examine the impact of genetic, psychological, and environmental factors on the health and well-being of mothers and children ^(20)^. The cohort is set in Bradford, a city in the north of England with high levels of socioeconomic deprivation and ethnic diversity. Pregnant women (n=12,453) were recruited between 2007 and 2010 while attending the Bradford Royal Infirmary for universal oral glucose tolerance testing at 26-28 weeks gestation. A semi-structured questionnaire on socio-demographic characteristics and health and lifestyle behaviours was self-completed by women and consent to routine linkage of mother and child data was obtained^(21)^. Multi-lingual research assistants provided support for non-English speaking mothers.

We used data from a sub-sample of the BiB cohort, the Born in Bradford 1000 longitudinal study (BiB1000, n=1,735), which was recruited between August 2008 and March 2009. BiB1000 aimed to examine early life factors such as diet, anthropometry, environmental, behavioural, and social factors that were hypothesised to be associated with the development of childhood obesity ^(22)^. All women recruited into the BiB birth cohort during this period were invited to take part in BiB1000. Ethical approval was granted by the Bradford Research Ethics Committee (ref: 07/H1302/112).

### Body weight

Women’s body mass index (BMI) was calculated as kg/m^2^ from measured weight (kg; Seca 2in1 scales, Harlow Healthcare Ltd, London, UK) at 24-months postpartum and measured height (m) from the baseline visit at approximately 26-28 weeks gestation. BMI status categories were then derived following convention: underweight (BMI<18.5), normal weight (18.5≤ BMI<25), overweight (25≤ BMI<30), and obese (BMI≥30)^(23)^.

### Sociodemographics and covariates

At baseline, women self-reported their ethnicity (white British; Pakistani; Other), country of birth (UK; Pakistan; Other), age (years), education (A-level equivalent or higher; maximum of 5 General Certificate of Secondary Education [GCSE], unknown, or foreign), parity, and how well they felt they were managing financially (struggling financially; not struggling financially). Women who identified as “Pakistani”, irrespective of country of birth, were categorised as “Pakistani-origin”. Homes were categorised into quintiles of national Index of Multiple Deprivation (IMD) using postcodes, with lower quintiles indicating higher deprivation. IMD is used as a measure of relative deprivation for small areas in England ^(24)^.

Cohabitation status (married; cohabiting with a partner; partner, not cohabiting; single) and physical activity were obtained from the 24-month postpartum questionnaire. Women’s reported physical activity at this time in minutes/week were classified into three categories based on UK government guidelines (sedentary [0 active minutes/week]; insufficiently active [≥1 active minute/week but ≤ 149 minutes/week]; sufficiently active [≥150 active minutes/week]) ^(25)^.

### Body weight perception

Body weight perception was assessed in the 24 months postpartum questionnaire using two methods, one posed as a question and one as a visual scale ^(26)^. In the former, women were asked a perceived weight status question (PWQ): “at this moment in time how would you describe yourself” and given a 7-item Likert-type scale ranging from “very overweight” through “just right” to “very underweight”. These items were condensed to represent the World Health Organization (WHO) cut-offs: underweight (“very underweight”, “moderately underweight”, “slightly underweight”); normal weight (“just right”); overweight (“slightly overweight”, “moderately overweight”); obese (“very overweight”). In the latter, women were provided with a Figure Rating Scale (FRS) which consists of a visual scale of nine female silhouettes that range from very thin (A) to very large (I)^(26)^. Women were asked to select the figure that looked most like them at that moment in time, with an option to select “don’t know”. Following cut-offs by other investigators, we categorised the FRS into: underweight (A-B); normal weight (C-D); overweight (E-G); obese (H-I) ^(27)^. Women who selected “don’t know” (n=10) were coded as missing.

Perception of body weight was considered accurate if women selected a perceived weight status category that matched their objectively measured weight category. We considered women with obesity who identified as overweight to accurately perceive their weight because it has been suggested that self-identification with a stigmatized group may be a driver in weight management behaviours ^(7)^. Women were considered to have overestimated their weight if they selected any weight category greater than their objectively measured weight category, i.e. if women were objectively measured to be underweight and perceived themselves as normal weight, overweight, or obese. Women were considered to have underestimated their weight if they selected a weight category lesser than their objectively measured weight category, i.e. if women were objectively measured to be overweight and perceived themselves as normal weight or underweight.

### Statistical analyses

All analyses were conducted in R version 3.3.3 (R Foundation for Statistical Computing, Vienna, Austria, 2017). We only considered women without missing data for the FRS or PWQ, with singleton pregnancies, and those of white British and Pakistani-origin as they were the dominant ethnic groups at the 24-month study visit (37% and 50%, respectively); the remaining ethnic groups would have too small and too ethnically diverse to contribute meaningfully to conclusions (total n=939). Only Pakistani-origin women (n=544) were analysed by country of birth as the majority (98.7%) of white British women were born in the UK. Participant characteristics were described by ethnic group and country of birth (UK-born; foreign-born). Sociodemographic characteristics, body weight data, and perceived body size by ethnicity and country of birth using the FRS and PWQ are presented as mean (standard deviation; SD) for continuous variables or percent for categorical variables. χ2 or Fisher’s exact test for categorical variables and t-test for continuous variables were used to assess differences in between white British and Pakistani-origin women, and between UK-born and foreign-born Pakistani-origin women.

Weighted kappa statistics were calculated to determine strength of agreement between objectively measured BMI categories and both measures of perceived weight status categories by ethnic group and by country of birth among Pakistani-origin women using the wkappa function from the *psy* package in R. Two-sided 95% confidence intervals (CI) were obtained through bootstrapping.

Logistic regression was used to assess the odds of underestimation compared to those who were accurate in their perception. We assessed odds of underestimation as under-detection of excess weight is more prevalent and has greater potential health implications ^(7)^. A directed acyclic graph (DAG) was drawn to assist in selecting covariates for adjustment (Supplementary figure 1) to estimate the total effect ^(28,29)^. Potential covariates included in the DAG were age, birth country, physical activity, IMD, education, subjective poverty, parity, and ethnicity. The DAG indicated no adjustment was needed in order to estimate ethnicity or birth country on misperception; however, we show analyses unadjusted and adjusted for age as age was the only variable in the DAG which was not in the causal pathway.

In sensitivity analyses, we explored the role of language on misperception by excluding women who did not complete the questionnaire in English (i.e. required language assistance). All white British women and English-speaking Pakistani-origin women were included (excluded from Pakistani-origin UK born: n=4 (1.9%); excluded from foreign-born: n=214 (63%)). We re-analysed the differences between categories of misperception, and logistic regression models by ethnicity and country of birth. Kappa agreement between the FRS and PWQ with perceived weight categories were only re-analysed for English-speaking Pakistani-origin women.

## Results

### Characteristics

White British and Pakistani-origin women were, on average, 27 years old (Table 1). Compared to white British women, Pakistani-origin women were more likely to be married (96% vs 43%), lived in the most deprived quintile of IMD (81% vs 51%), were less likely to be sufficiently active (27% vs 55%), and were less likely to be obese (23% vs 30%).

**Table 1.**
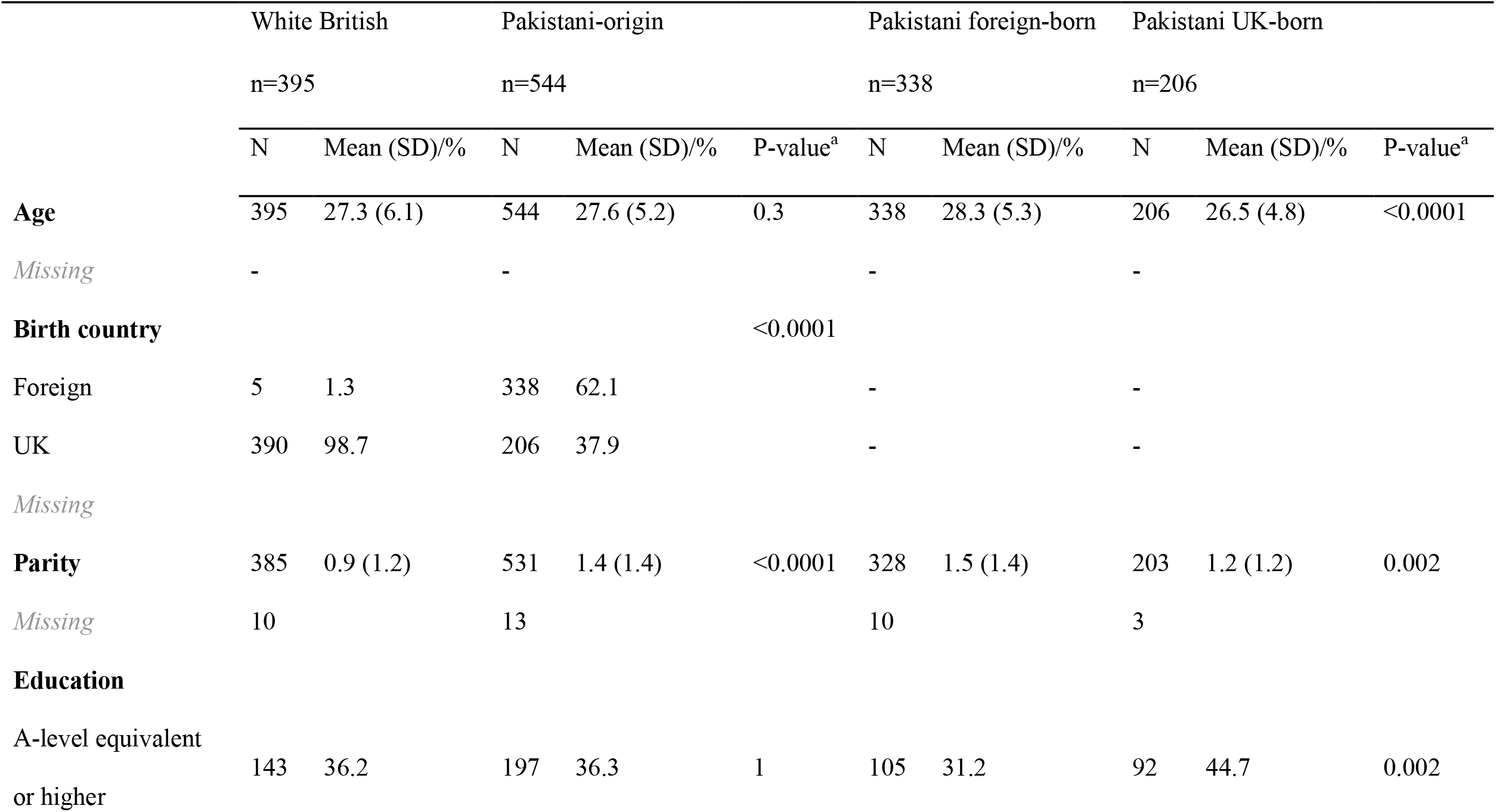
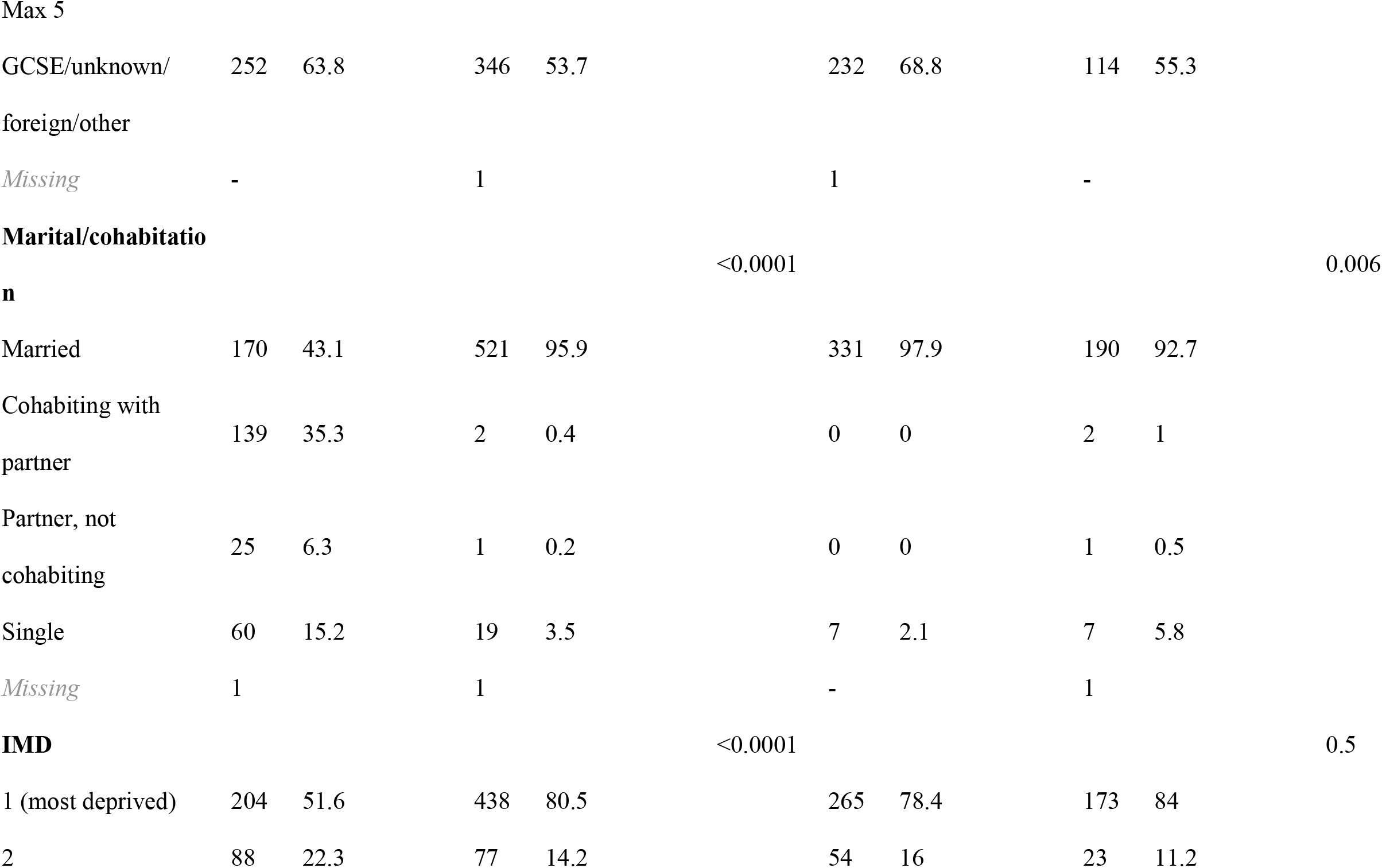
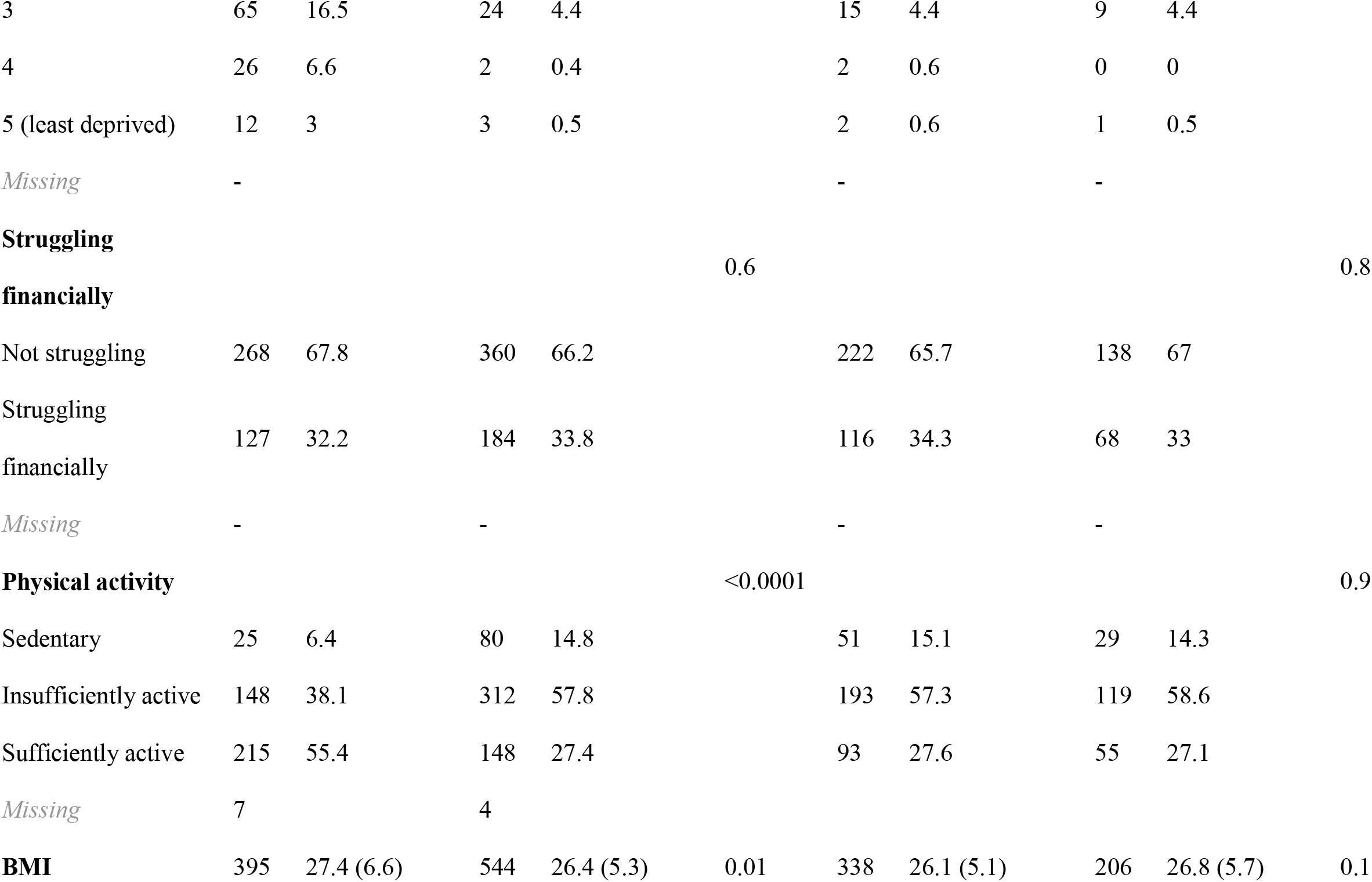
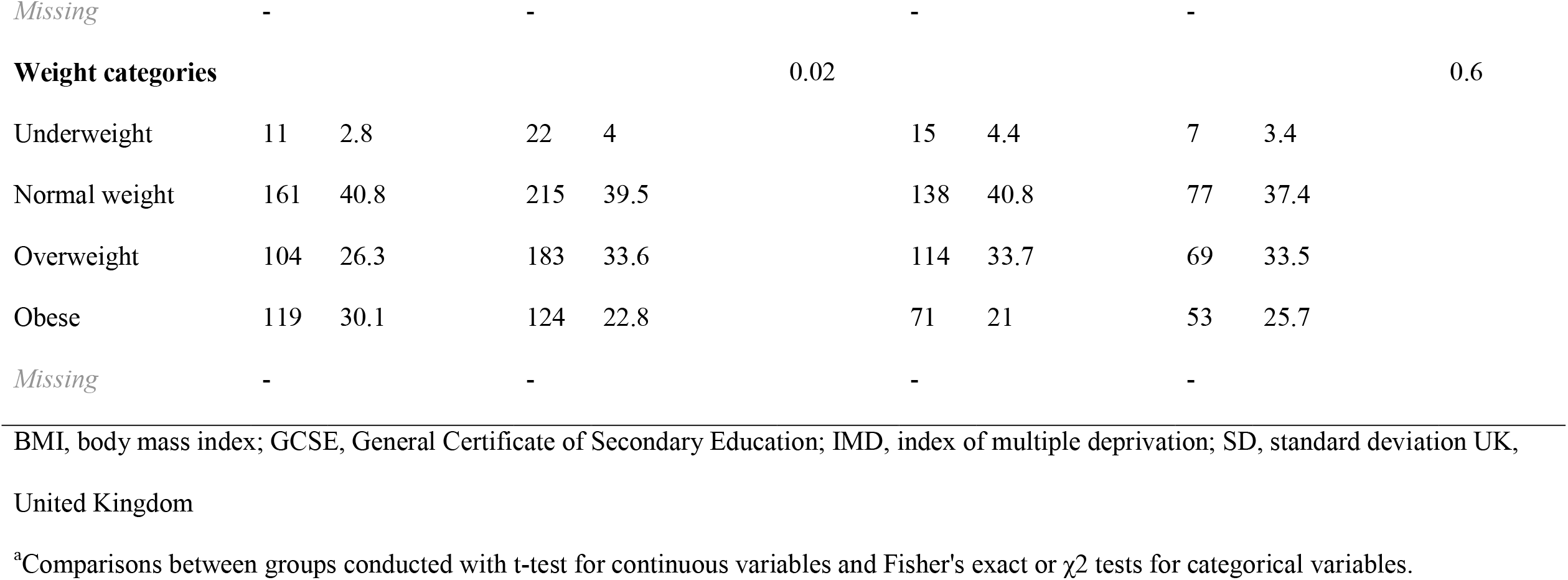
Characteristics of women in the Born in Bradford cohort by ethnicity and by country of birth.

The majority (62%) of Pakistani-origin women were not born in the UK. Compared to Pakistani women who were born in the UK, foreign-born women were older (mean [SD]: 28.3 [5.3] vs 26.5 [4.8] years), had more children (1.5 [1.4] vs 1.2 [1.3]), were less likely to have higher levels of education (31% vs 45%), and more likely to be married (98% vs 93%).

### Weight misperception

White British women were more accurate in their body weight perception than Pakistani-origin women, who were more likely to underestimate their body weight using both the FRS and PWQ (Figure 1a). Fourteen percent of Pakistani-origin women underestimated their body weight compared to 6% of white British women using the FRS. With the PWQ, 18% of Pakistani-origin women and 10% of white British women underestimated their body weight.

**Figure 1.**
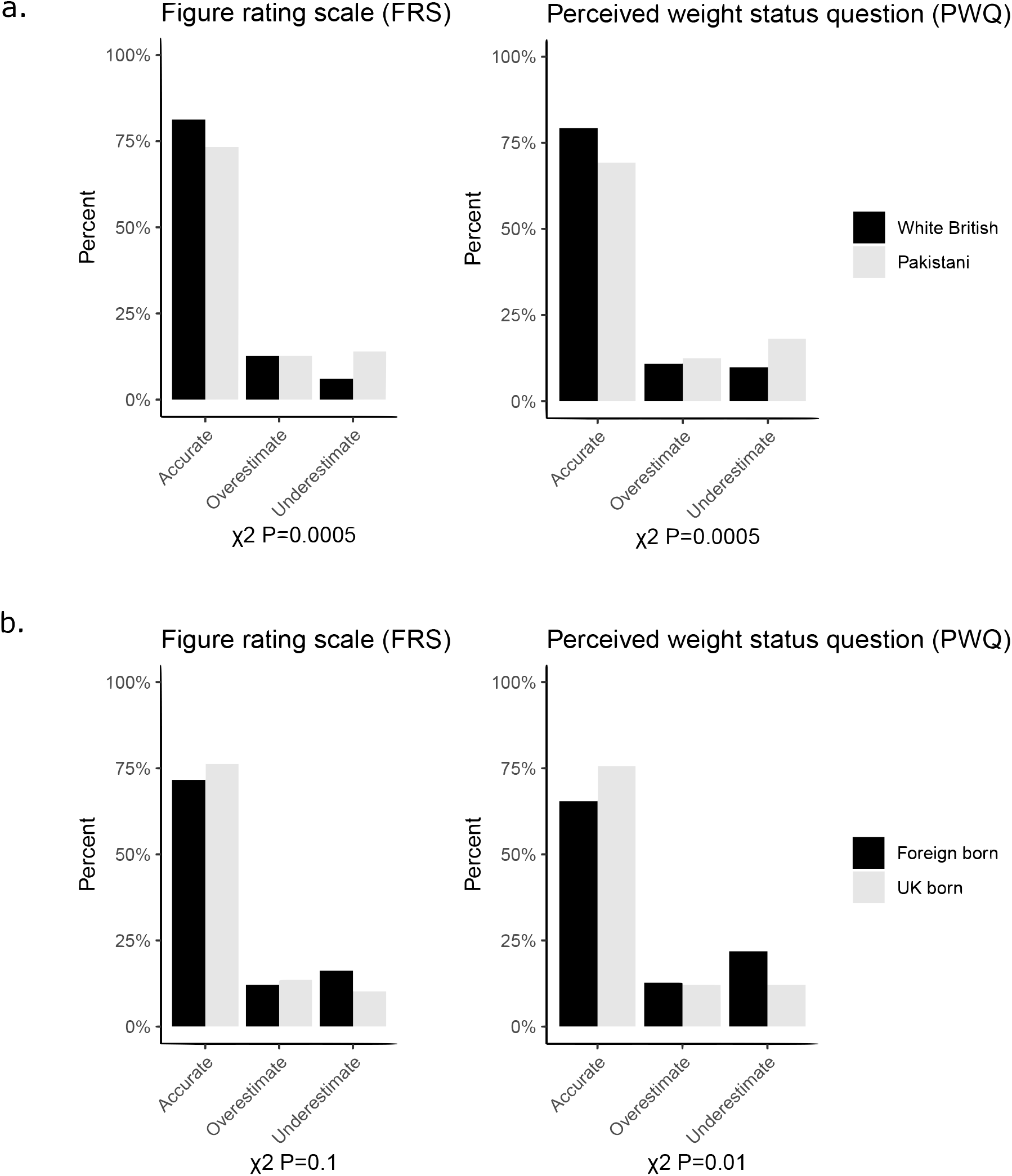
Accuracy of body weight perception using the figure rating scale (FRS) and perceived weight status question (PWQ) by (a) ethnicity and (b) country of birth among Pakistani-origin women.

Kappa statistic agreement between objective and perceived body weight were similar for white British women using both methods (FRS: κ=0.61; 95% CI=0.56, 0.65; PWQ: κ=0.61; 95% CI=0.55, 0.68). For Pakistani-origin women, agreement was greater with the FRS (κ=0.58; 95% CI=0.52, 0.63) than the PWQ (κ=0.47; 95% CI=0.40, 0.54).

Among Pakistani-origin women, those born in the UK were more accurate in their body weight perception compared to those foreign-born (Figure 1b). Ten percent of Pakistani-origin women born in the UK underestimated their body weight compared to 16% of women foreign-born using the FRS. With the PWQ, 12% of Pakistani-origin women born in the UK and 22% of those foreign-born underestimated their body weight.

Kappa statistic agreement between objective and perceived body weight were similar for Pakistani-origin women born in the UK with both methods (FRS: κ=0.57; 95% CI=0.47, 0.65; PWQ: (κ=0.55; 95% CI=0.47, 0.53). For Pakistani-origin women born outside the UK, agreement was greater with the FRS (κ=0.59; 95% CI=0.52, 0.65) than the PWQ (κ=0.42; 95% CI=0.32, 0.53).

### Odds of underestimation

Compared to white British women, Pakistani-origin women were more likely to underestimate their body weight; this was greater when using the FRS (154% more likely) than the PWQ (111% more likely) (Table 2). Among Pakistani-origin women, foreign-born women were more likely to underestimate their body weight when assessed using the PWQ (105% more likely compared to those born in the UK).

**Table 2.**
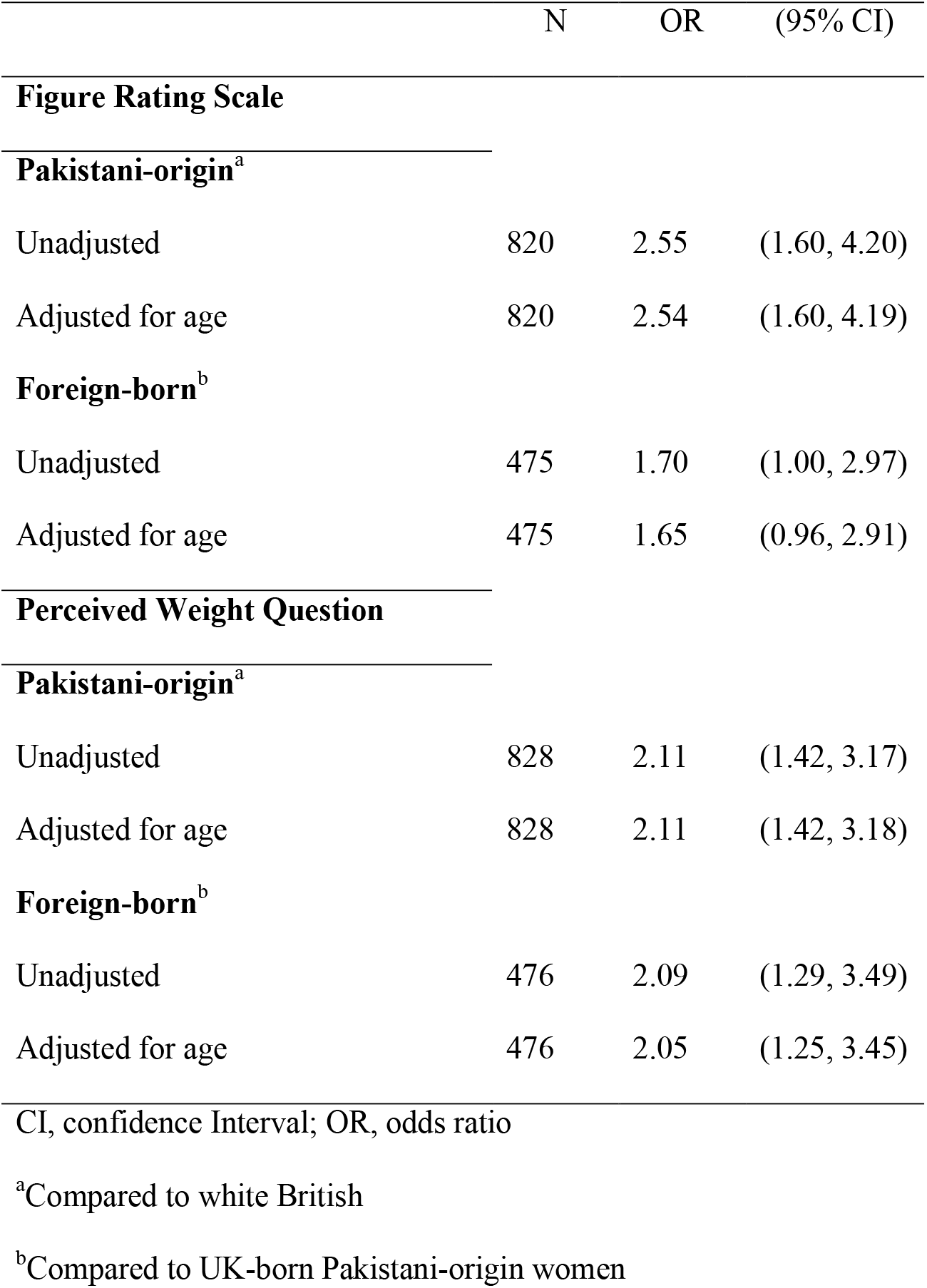
Odds of underestimation by ethnicity and country of birth among Pakistani-origin women.

|  | N | OR | (95% CI) |
| --- | --- | --- | --- |
| **Figure Rating Scale** |  |  |  |
| **Pakistani-origin<sup>a</sup>** |  |  |  |
| Unadjusted | 820 | 2.55 | (1.60, 4.20) |
| Adjusted for age | 820 | 2.54 | (1.60, 4.19) |
| **Foreign-born<sup>b</sup>** |  |  |  |
| Unadjusted | 475 | 1.70 | (1.00, 2.97) |
| Adjusted for age | 475 | 1.65 | (0.96, 2.91) |
| **Perceived Weight Question** |  |  |  |
| **Pakistani-origin<sup>a</sup>** |  |  |  |
| Unadjusted | 828 | 2.11 | (1.42, 3.17) |
| Adjusted for age | 828 | 2.11 | (1.42, 3.18) |
| **Foreign-born<sup>b</sup>** |  |  |  |
| Unadjusted | 476 | 2.09 | (1.29, 3.49) |
| Adjusted for age | 476 | 2.05 | (1.25, 3.45) |

CI, confidence Interval; OR, odds ratio
<sup>a</sup>Compared to white British
<sup>b</sup>Compared to UK-born Pakistani-origin women

### Sensitivity analysis

Among women with English proficiency, Pakistani-origin women were less accurate in perceiving their body weight compared to white British women using the FRS (Supplementary figure 2). In logistic regression models, Pakistani-origin women were 127% more likely to underestimate their body weight category compared to white British women when using the FRS (Supplementary table 1). We did not find differences in misperception or odds of underestimation by country of birth.

The FRS was similar to, or better than, the PWQ for agreement between objective and perceived weight status categories for English-proficient Pakistani-origin women overall (FRS: κ=0.57; 95% CI: 0.51, 0.64; PWQ: κ=0.53, 0.44, 0.62). Agreement was similar using either method for those born in the UK (FRS: κ=0.57; 95% CI=0.47, 0.66; PWQ: κ=0.56; 95% CI=0.47, 0.65) while the FRS showed greater agreement for those foreign-born (FRS: κ=0.57; 95% 0.45, 0.66; PWQ: κ=0.45; 95% CI=0.27, 0.61).

## Discussion

We observed that accuracy of weight perception differed by ethnicity but not country of birth and that the method of weight perception assessment can influence the degree of misperception. Agreement between objective and subjective measurements using Kappa statistics found the FRS a more accurate tool for assessing weight perception, particularly among foreign-born women. We found white British women were more likely to accurately perceive their body weight and Pakistani-origin women were more likely to underestimate their body weight using both assessment methods. However, among Pakistani-origin women, differences in misperception by country of birth were more pronounced for those foreign-born and only observed when self-assessment was carried out through the PWQ; these relationships disappeared when language was taken into account by excluding non-English speaking women. Taken together, these results suggest that, while ethnic differences in accuracy of body weight perception exists, using a PWQ rather than a visual scale to assess body weight perception can distort the number of women who underestimate their body weight.

Misperception of body weight among South Asian women has been reported by others, though whether differences in migration status to the UK have not been previously explored. Similar to other studies, we found that Pakistani-origin women were more likely to underestimate their body weight compared to white British women. In a study in West London, South Asian women were less likely to consider themselves as overweight, when they were objectively measured as overweight, compared to white European women (62% vs 93%) ^(30)^. Similarly, another study in the UK found 43.5% of South Asian women perceived themselves as being “about the right weight” when they were objectively measured as overweight, compared to 6.3% of European women ^(31)^.

In our study, Pakistani-origin women born outside the UK had greater levels of misperception, particularly underestimation, compared to Pakistani-origin women born within the UK. Underestimation of body weight also appears among adults within Pakistan, the birth country of all our foreign-born Pakistani-origin women. Bhanji et al. (2011) found only 27% of individuals with obesity accurately perceived their obesity while the same number of those with obesity thought their weight was “just right” and almost half of those with overweight considered their weight to be “just right” ^(32)^. The differences in the level of misperception observed between Pakistani-origin women born in the UK or foreign-born may be due to different norms and cultural beliefs about body size ^(13,14)^. Bush et al. (2001) examined the attitudes of UK first generation and migrant South Asian women towards body size and observed that migrant women had greater BMI, waist circumference, and waist/hip ratio and associated positive health with larger body sizes. In contrast, those who were British-born tended towards more Western ideals against larger body sizes ^(14)^. Migrant South Asian women associated “prestige” (securing a good job; marriage) with thinner silhouettes, but were more likely to equate larger body sizes (equivalent to BMI≥28) with being more likely to eat healthy food, have healthy children, and be healthier. The authors hypothesised that migrant women to the UK were only beginning to perceive excess body weight as being a potential issue, leading to the inconsistency in their perception of different silhouette sizes with health and life outcomes. Excess weight may also not be a cause for concern until it presents health problems; a study of South Asian Americans reported that individuals with overweight and obesity only perceived their excess bodyweight as problematic when it resulted in physical limitations such as joint aches, inability to exercise, and shortness of breath ^(15)^.

This is the first study to assess differences in body weight perception by migration through country of birth in the UK. We found that foreign-born Pakistani-origin women were more likely to underestimate their body weight compared to Pakistani-origin women born in the UK but only when using the PWQ. However, when we excluded non-English speaking women, there were no differences in misperception by country of birth. This exclusion meant over 60% of foreign-born Pakistani-origin women needed translation assistance to complete the questionnaire. Difficulties in translating the different weight categories or their meaning for the PWQ may have led to misclassification and resulted in spurious differences in levels of misperception when non-English speaking women were included in the analysis. The perceived weight status question was worded using clinical terms rather than the “plain English” terms such as “thin” and “fat” used by other studies ^(12)^. Words and concepts such as “overweight” may be perceived to mean different things in different cultures and indicates that the measurement tool used may have a role in influencing the degree of accurate body weight perception ^(33,34)^. As results did not change between Pakistani-origin women by country of birth using the FRS, it can be hypothesised that the use of a visual scale is more easily understood universally. The kappa agreement between the FRS and PWQ and objective weight categories, while overlapping in confidence intervals, also suggest that the FRS is a better tool overall. These results suggest that a visual scale would be the most appropriate tool as it doesn’t rely on clinical or stigmatizing terms and can be considered more generalizable and transferable.

There are several limitations to this study. The women in this study are participants in a birth cohort study and live in a deprived city in the north of England. This limits the generalizability of the findings. However, UK studies of misperception among white British and South Asian women have similar findings between groups and studies from the US have also noted differences in perception by ethnic group ^(11,13,30,31)^. Over 60% of our Pakistani-origin women were not able to complete the questionnaire in English; their exclusion from our sensitivity analyses reduced our sample size and power to explore the potential role of language in accurate self-assessment. However, this analysis was exploratory and the diversity of language in this cohort adds to its unique ability to examine this concept. We also did not adjust for a range of covariables; it is possible that our DAG did not accurately depict the relationships between variables and differences observed in misperception may be due to factors other than ethnicity and country of birth. The PWQ and FRS were asked sequentially in the questionnaire, which could have made women more aware of their body size. The Stunkard figure rating scale, while widely used, has a number of methodological concerns such as the number of figures shown and the inconsistency in figure sizes when “scaling up” and other scales may be more appropriate ^(35,36)^. The study visit procedure had the mother complete the questionnaire before being weighed but allowed for the mother to be weighed before completing the questionnaire, depending on the child’s behaviour; this could potentially influence the response on self-perception but this occurrence is likely random and its consequence would be to mask the true effect.

In conclusion, we observed differences in weight misperception by ethnicity with greater underestimation among Pakistani-origin women. The method used to assess perceived body weight status is important; self-assessment using visual scales more strongly agrees with objective weight status and may be understood more consistently across populations. Studies looking to measure perceived weight status should employ visual scales where possible.

## Data Availability

The data underlying this article will be shared on reasonable request to the corresponding author.

## Acknowledgements

Born in Bradford is only possible because of the enthusiasm and commitment of the Children and Parents in BiB. We are grateful to all the participants, practitioners, and researchers who have made Born in Bradford happen.

## Data availability

The data underlying this article will be shared on reasonable request to the corresponding author.

