## Supplementary file 1 for "Ethnicity, migration, and weight misperception: a Born in Bradford study"

Supplementary Figure 1. Directed acyclic graph (DAG) drawn to depict the relationship between ethnicity and birth country with covariates and misperception of body weight.

**Birth country      misperception**

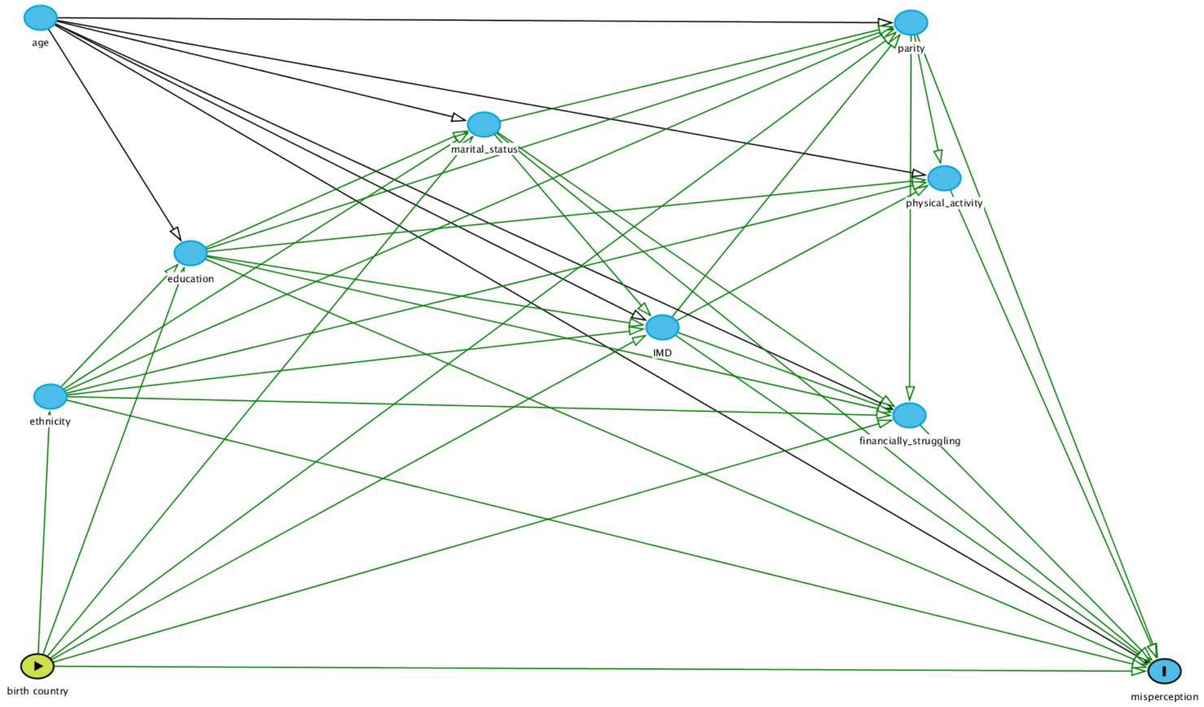


**Ethnicity      misperception**

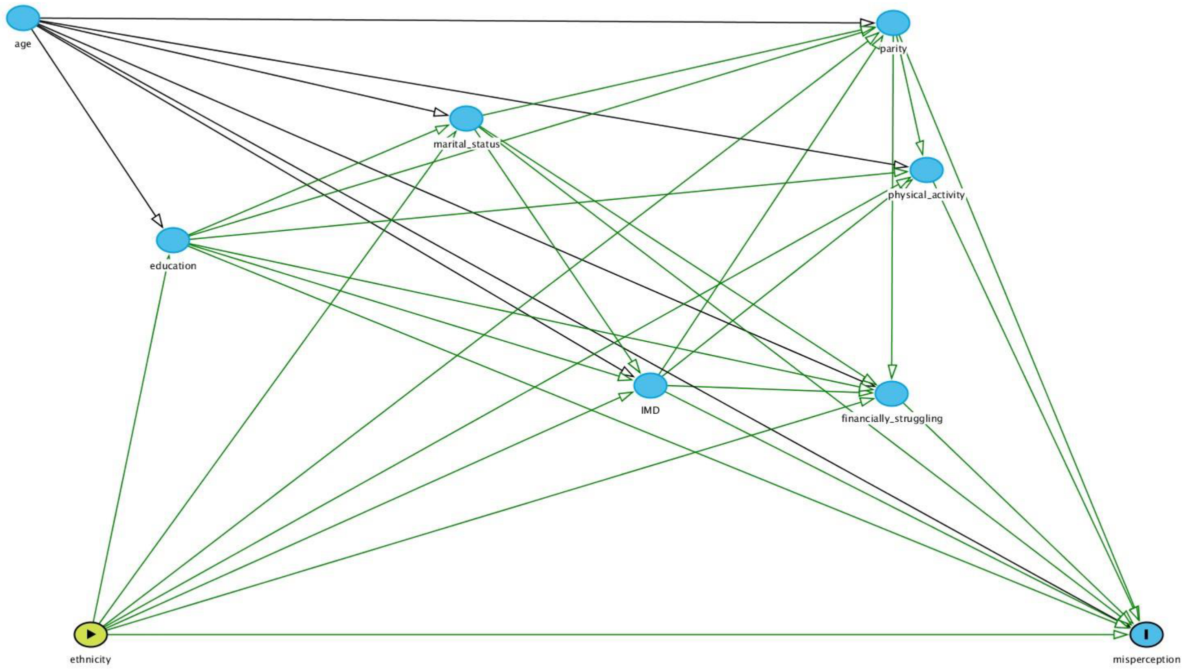
