## Supplementary file 2 for "Ethnicity, migration, and weight misperception: a Born in Bradford study"

Supplementary figure 2. Accuracy of body weight perception using the figure rating scale (FRS)and perceived weight status question (PWQ) among English-speaking women by (a) ethnicity and (b) country of birth among Pakistani-origin women.

a.

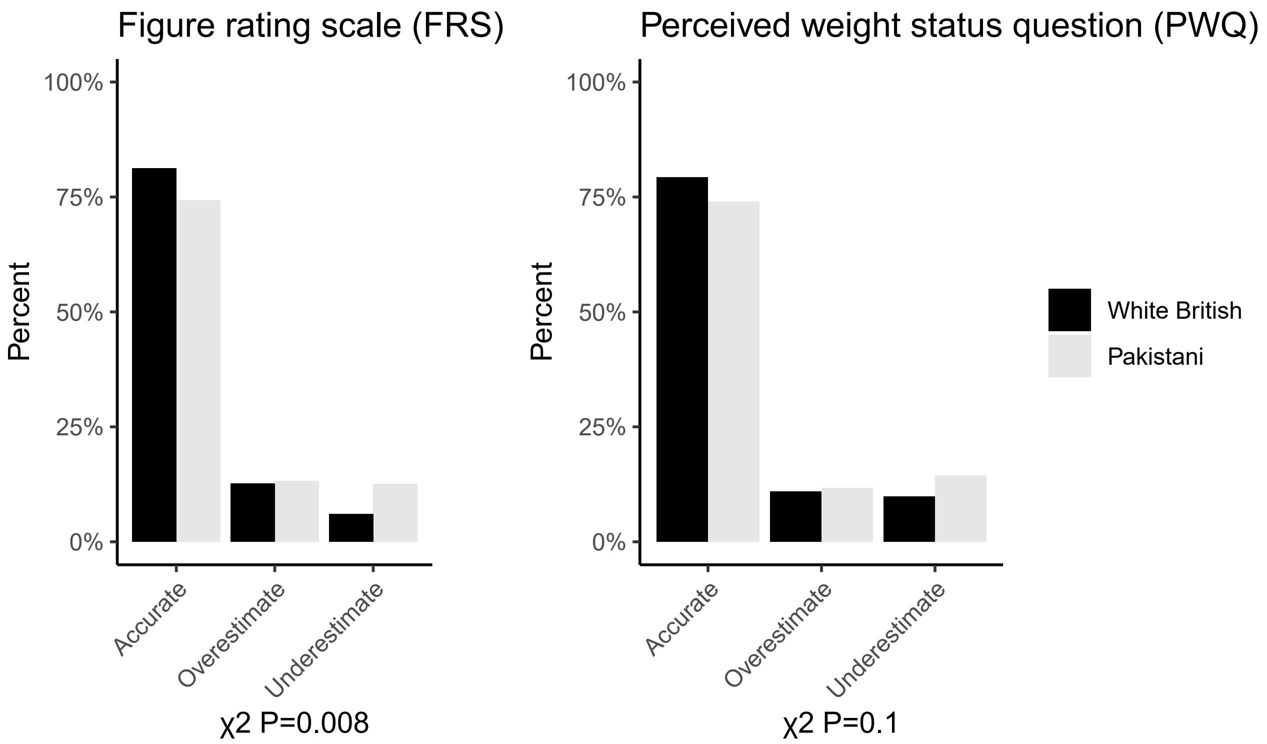


b.

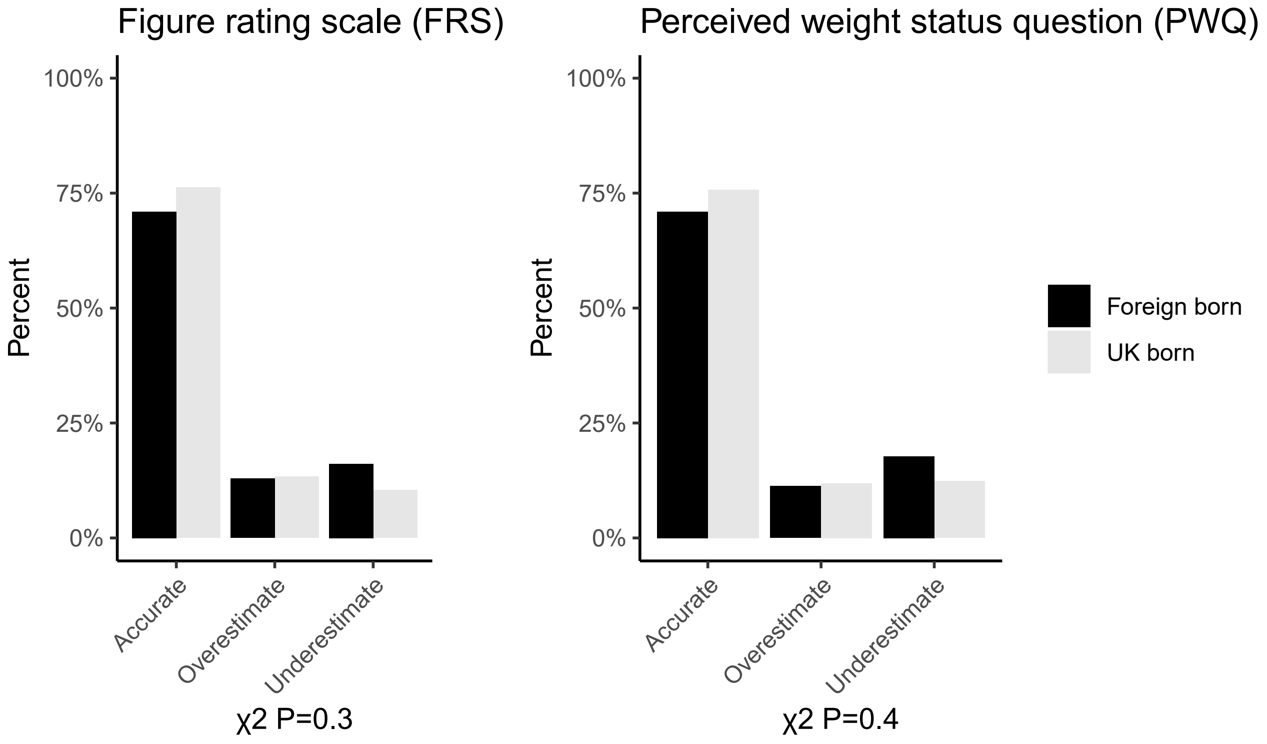
